# HIV STATUS AND ITS ASSOCIATED FACTORS OF CHILDREN BORN TO HIV POSITIVE MOTHER ON ART AT GAMBELLA PUBLIC HOSPITALS, SOUTH WEST ETHIOPIA, 2024

**DOI:** 10.1101/2025.08.01.25332598

**Authors:** Abel Felege, Daniel Girma, Haymanot Berihun

## Abstract

**Background:** In the 21 Global Plan priority countries in SSA, including Ethiopia, 110,000 children are newly infected with HIV annually. Most pediatric infections occur during breastfeeding through vertical transmission, with a transmission risk of 15%-45%, which can be reduced to 5% with effective interventions. Despite declining MTCT rates in Ethiopia, recent studies still report significant infection rates in infants.

**Objective:** HIV status and its associated factors in children born to HIV-positive mothers on art at Gambella Public Hospitals, Southwest Ethiopia, 2024.

**Methods:** This was an institution-based cross-sectional study in which three public hospitals in the Gmbella region were included. The sample size for this study was determined via the single population proportion formula and EPIINFO software, resulting in 250 participants after accounting for a 10% nonresponse rate. Three hospitals were randomly selected, and proportional allocation on the basis of the average two-month ART clinic visits was used to determine the sample sizes, ensuring a comprehensive representation. The data were collected via the interview method. The data were entered into Epi-Data 4.6.0.2 and exported to SPSS version 26.00 for clearing and analysis, and logistic regression analyses were employed to identify factors associated with HIV status. Using 95% CIs, variables with a p value <0.05 were identified as statistically significant factors.

**Results:** This study revealed that 7% of children born to HIV-positive mothers were also infected with HIV. Mothers aged 30–39 years (AOR=8.7), place of residence (rural) (AOR=6.7), positive results for syphilis (AOR=19.6), and unplanned pregnancies (AOR=10.4) were significantly associated with the outcome variable.

**Conclusion:** This study revealed a 7% HIV infection rate among children born to HIV-positive mothers in Gambella, with maternal age, rural residence, syphilis-positive mothers, and unplanned pregnancies associated with HIV infection.

## Introduction

It is estimated that 39 million people worldwide are living with HIV and acquired immunodeficiency disease syndrome (AIDS), with more than two-thirds residing in sub-Saharan Africa (1). Human immunodeficiency virus (HIV) can be transmitted from HIV-positive mothers to their babies during pregnancy, delivery, and breastfeeding, accounting for the majority of infections in children (2, 3). The rate of mother‒to-child transmission (MTCT) of HIV in untreated pregnant women ranges from 15% to 45%. However, antiretroviral therapy (ART), postexposure prophylaxis, and prevention of mother‒child transmission (PMTCT) services have significantly reduced the MTCT rate to less than 5% (4, 5). Virological screening for exposed infants at six weeks of age, or as early as possible, plays a crucial role in PMTCT (6).

In 2019, 1.8 million children under 15 years of age were living with HIV, with 15,000 newly infected children and 95,000 AIDS-related deaths among children. Globally, approximately 4,500 new HIV infections occur daily, with 59% of these occurring in sub-Saharan Africa (1). Efforts to reduce new HIV infections among children have been made at both the global and national levels. The World Health Organization (WHO) promotes a comprehensive PMTCT approach with four principles: primary prevention of HIV among reproductive-age women, prevention of unintended pregnancies among women living with HIV, prevention of MTCT, and provision of treatment and support to mothers and children living with HIV (3).

Since 2012, the WHO has recommended Option B+ for PMTCT, which involves providing lifelong ART to all pregnant and breastfeeding women regardless of their CD4 count (7). This approach has significantly reduced MTCT rates to less than 2% and decreased the degree of sexual transmission of HIV to uninfected partners (8, 9). ART during and after pregnancy has averted an estimated 1.3 million new HIV infections among children (9). Studies have shown that with effective intervention, the risk of MTCT can be reduced to as low as 1% to 5% at six months (10, 11).

A systematic review and meta-analysis of pooled data reported from different parts of Ethiopia indicated that 11.4% and 9.9% of infants are HIV-infected through MTCT (12, 13). Conducting research on HIV infection in children is crucial for several compelling reasons, given the profound implications for individual health, public health, and societal well-being. Research is essential in refining and improving PMTCT strategies to reduce this risk. Studies have shown that early diagnosis and initiation of ART significantly improve survival rates and health outcomes for HIV-positive children. Treating HIV in children involves significant healthcare resources. Research into these areas can inform policies and programs to support social integration and psychological well-being.

This study sought to investigate the HIV status of children born to HIV-positive mothers and identify the factors associated with MTCT among HIV-positive mothers in Gambella Public Hospitals.

HIV/AIDS remains a significant public health issue, particularly in sub-Saharan Africa. Despite advancements in prevention and treatment, MTCT remains a critical challenge (14). In 2022, UNAIDS reported that approximately 1.8 million children (aged 0--14) were living with HIV globally, with approximately 150,000 new infections among children annually (1). The SSA region bears the highest burden of HIV, including MTCT, with limited access to prenatal care and ART in resource-limited settings, hindering efforts to prevent MTCT (3).

The impact of HIV on children is profound, affecting their health, growth, and neurological development and increasing their susceptibility to infections. Without treatment, approximately 50% of children living with HIV die before their second birthday. In 2022, approximately 99,000 children died from AIDS-related causes (4, 6, 15). The cost of lifelong ART, medical check-ups, and treatment for opportunistic infections is substantial (6). Families with HIV-infected children often face increased caregiving responsibilities, leading to a loss of income and productivity and high out-of-pocket expenses for medical care and special nutritional needs (4, 6, 15)

Efforts to minimize HIV infection among children have involved a combination of medical, social, and policy interventions. Pregnant women living with HIV are provided with ART to reduce viral load and transmission risk. The WHO recommends lifelong ART for all pregnant and breastfeeding women living with HIV (3).. Early diagnosis and antiretroviral prophylaxis for HIV-exposed infants are crucial. Elective cesarean delivery is recommended for women with high viral loads (16).

Prevention of MTCT (PMTCT) programs are integrated into maternal and child health services. Awareness campaigns, community involvement, and training programs for healthcare providers play essential roles in reducing stigma and improving the management of HIV in pregnant women and children (3, 17–19).

The factors associated with children being HIV positive are multifaceted and include maternal viral load, mixed feeding practices, and economic and social barriers. Late initiation or inconsistent adherence to ART increases transmission risk. Conflict and displacement also heighten vulnerability to HIV infection (4, 17).

In Ethiopia, 92% of pregnant women living with HIV had access to ART in 2020, leading to a significant decline in MTCT incidence (14). However, despite the overall decline, some regions continue to report considerable infection rates among infants born to HIV-positive mothers (14, 20–22)

### Conceptual framework

Illustrated in Figure 1.

**Figure 1.**
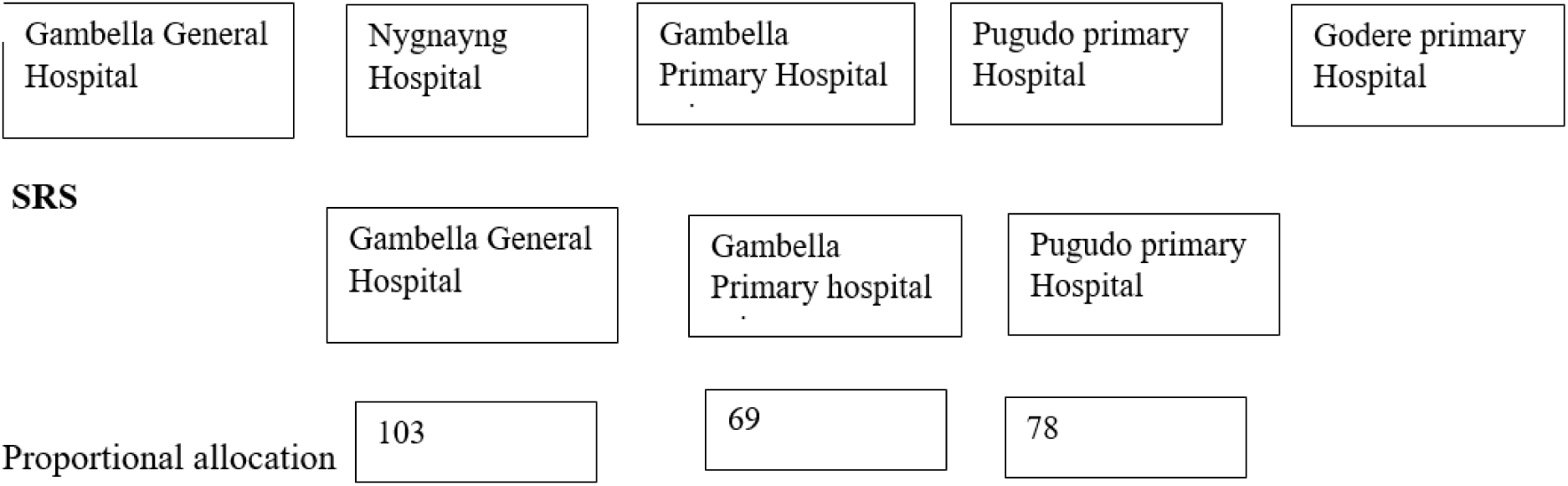
Sampling Procedure for HIV Status and Its Associated Factors of Children Born to HIV Positive Mothers on ART at Gambella Public Hospitals, Southwest Ethiopia, 2024

### General objective

To assess HIV status and its associated factors among children born to HIV-positive mothers on ART at Gambella public hospitals, Southwest Ethiopia, 2024.

### Specific objective

To determine the magnitude of HV infection among children born to HIV-positive mothers on ART at Gambella public hospitals, southwestern Ethiopia, 2024.

To identify factors associated with HIV among children born to HIV-positive mothers on ART at Gambella public hospitals in southwestern Ethiopia, 2024.

## Methods and Materials

### Study setting, design and period

The facility-based cross-sectional study design was conducted from June 15 to August 15, 2024. The Gambella region is located 670 km away from Addis Ababa in southwestern Ethiopia. Gambella is bordered by South Sudan to the west and the Ethiopian regions of Oromia and the Southern Nations, Nationalities, and Peoples’ Region (SNNPR) to the north and east, respectively. Gambella has 5 hospitals, 29 health centers, and 142 health posts. According to figures from the Central Statistical Agency (CSA) of Ethiopia, the Gambella region has an estimated total population of 483,097, consisting of 186,526 males and 171,985 females. Approximately 74.8% of the population was estimated to be rural, whereas 25.2% were urban dwellers. The research was implemented in public hospitals in the Gambella regional state of Ethiopia

### Population

The source population is all HIV-exposed children enrolled in the PMTCT clinics of Gambella public hospitals in southwestern Ethiopia. Our study population included children born to HIV-positive mothers on ART who had HIV results in the study setting during the study period.

### Inclusion and Exclusion Criteria

Inclusion criteriaChildren with DBS/PCR results during the study period and an age less than or equal to 18 months were included in our study, and children less than 45 days of age, infants with an unknown HIV status, mothers not on ART, infants who did not have a living mother and those unwilling to participate were excluded from our study.

### Sample size determination and sampling procedure

The sample size for this study was determined via the single population proportion formula, considering that 10.1% of the children in the Amhara region were HIV positive (22). The maximum sample size was calculated with a 5% margin of error and a 95% confidence interval, resulting in a sample size of 140 for the first objective. Using EPIINFO software for the second objective, the study considered various characteristics with 95% CIs and 80% power, leading to a maximum sample size of 250 after accounting for a 10% nonresponse rate, as illustrated in Table 1.

**Table 1.**
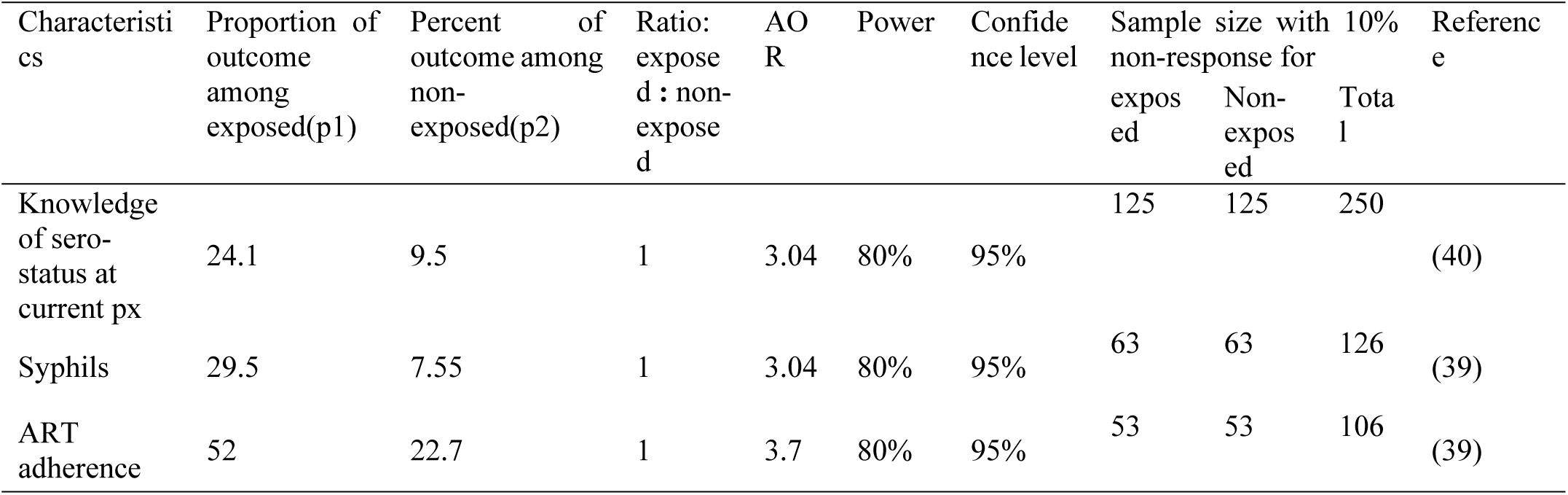
Sample Size Calculation for HIV Status and Its Associated Factors of Children Born to HIV Positive Mothers on ART at Gambella Public Hospitals, Southwest Ethiopia, 2024.

Three hospitals (Gambella General Hospital, Gambella Primary Hospital, and Pugnudo) were randomly selected, and proportional allocation was based on the average two-month ART clinic visit for children. The total average number of visits was 290, and individuals were selected randomly via simple random sampling on the basis of proportional allocation (Figure 2).

**Figure 2.**
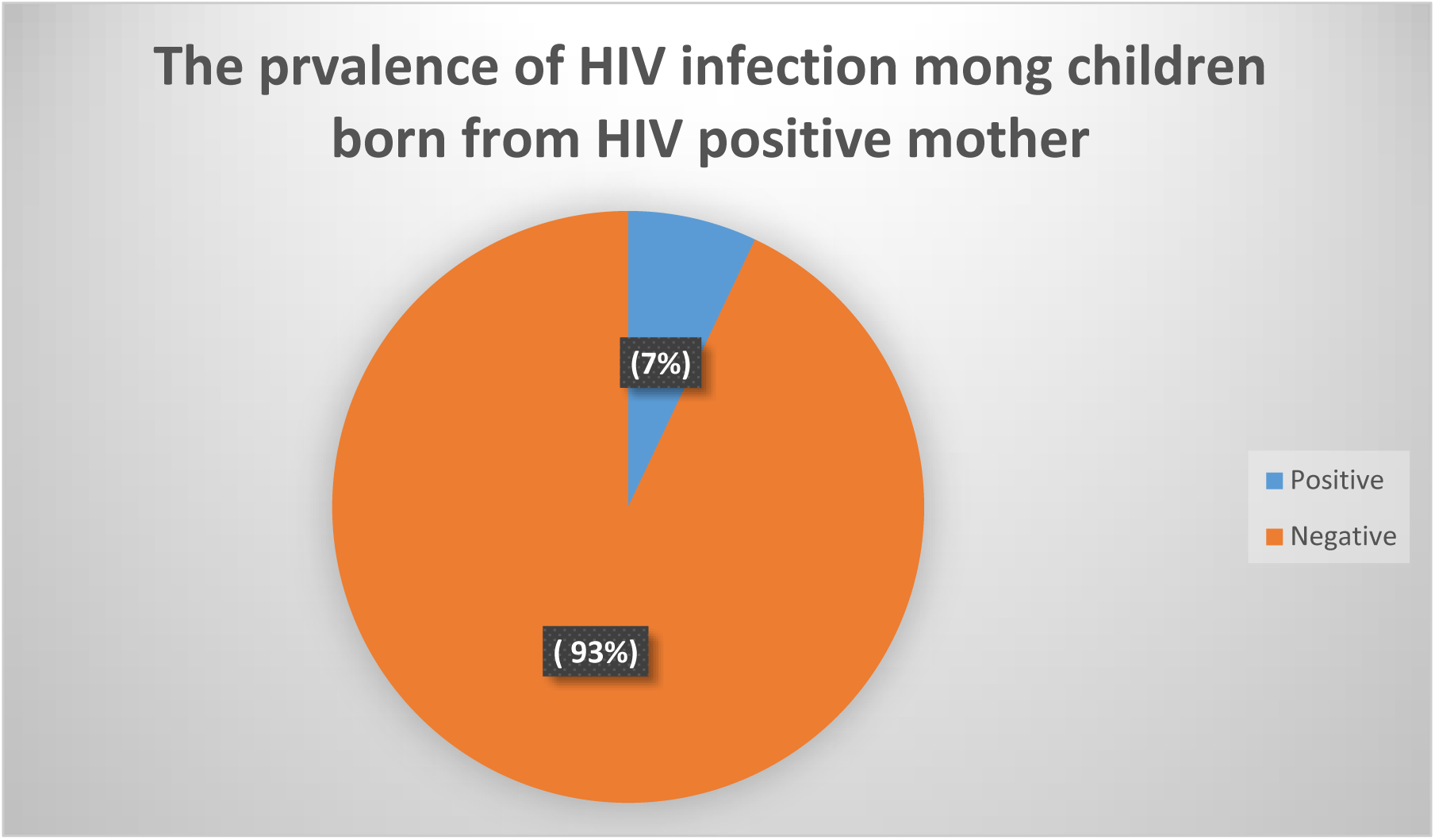
HIV Status of Children Born to HIV Positive Mothers on ART at Gambella Public Hospitals, Southwest Ethiopia, 2024

### Study variables

The dependent variable is the HIV status of HIV-exposed children. The independent variables include sociodemographic factors such as age, residency, education level, and occupation. Obstetric-related factors such as gravidity, parity, condition of pregnancy, place of delivery, and ANC are also considered. Comorbidities and ART-related characteristics include syphilis, nutritional status, adherence to ART, time of initiation, and duration of ART. Social factors include mother-to-mother support, partner involvement, and health facility-related characteristics, such as the distance of the health facility and the availability of treatment and prophylaxis. Child-related characteristics included NVP and AZT duration, time of ART initiation, type of feeding, sex, and weight at birth.

### Data collection procedure

A structured questionnaire adapted from national HIV-exposed infant protocols and various research studies on similar topics was used for this study. Nurses working in the ART unit conducted face‒to-face interviews with the study participants and carried out MUAC measurements after receiving two days of training. Two clinical nurses supervised the data collection process. The pretested structured questionnaire, initially prepared in English and translated into the local language, was employed to collect data. The participants were requested to complete the questionnaire after providing informed consent and screening for exclusion criteria. Primary data and registered information were collected from caregivers of the children in the selected health facilities via the interview method from June 15 to August 15, 2024, over a two-month duration. For each facility, one nurse was assigned to collect data. During data collection, supervision was carried out, and the collected data were checked daily. The data collectors were supervised, and the questionnaire was checked for completeness and accuracy to ensure validity. Any problems that arose during the data collection process were addressed with appropriate intervention by the principal investigator. Training for the data collectors and the supervisor was conducted on the planned date, and all the instruments were pretested before the actual data collection date.

### Data Quality Control

The questionnaire was prepared in English with an option for the local language, Amharic. The translation was performed by qualified professionals. A one-day training session was conducted for the data collectors on how to fill out the questionnaire and manage the overall data collection process before the actual data collection period. A pretest was conducted with 5% of the sample size at the Bonga Health Center. During data collection, the data were checked daily for completeness and consistency.

### Operational and term definitions

Definitive HIV test results HIV test result identified with DNA/PCR prior to 18 months of age, or by rapid antibody test after 18 months of age and 6 weeks of cessation of breastfeeding (19)

Mothers on PMTCT Mothers who have taken ART for prevention of MTCT of HIV either during pregnancy or child birth and delivery or during breastfeeding (19)

Nutritional status of the mother, as measured via mid-upper arm circumference (MUAC): if > 22 cm = not malnourished, ≤ 22 cm = malnourished. (19)

HIV-positive mothers who pass through PMTCT services and participate in at least one regular meeting of PMTCT.

Good adherence= if the number of missed doses within 60 days is three or fewer doses (19)

Fair adherence= if the number of missed doses within 60 days is four to eight(19)

Poor adherence= if the number of missed doses within 60 days is nine or more doses(19)

Exclusive breastfeeding Give only breast milk during the first six months of life and no other drinks or food, not even water, except for syrup containing vitamins and minerals.(23)

Breast milk is mixed with nonhuman milk and other fluids or solids(23)

Positive DBS= positive result from record that was done in hospital

Negative DBS= negative result from record that was done in hospital

### Ethics statement

This study was approved by the institutional review board (IRB) of the Ethical Review Committee of the College of Public Health of Mettu University. The study was conducted following the relevant guidelines, regulations, and principles of the Helsinki Declaration. Additionally, permission to conduct the study was obtained from the medical directors of three hospitals. Written consent was obtained from each of the study participants after their caregivers provided information about the nature and objective of the study. The participants were informed that they had the right not to participate in the study or could withdraw at any point without any repercussions on the quality of care they received. The confidentiality of the information was maintained by avoiding any personal identifiers, such as the patient’s name, on the questionnaires during the data collection. The information obtained from the participants was used solely for the study and was kept confidential to avoid any harm. Finally, the recorded data were kept safe by locking them in a secure location, accessible only by the principal investigator.

### Data analysis

The data were cleaned and entered into Epidata 4.6.0.2 and exported to SPSS version 26 for analysis. Summary statistics of the means and percentages were used to describe the study. Fitted bivariate logistic regression models were used to assess the associations between each of the study outcomes and the different potential risk factors. Then, multivariable logistic models were fitted to identify independent factors. For the multivariable regression modeling, the covariates were included in a model that was selected on the basis of their bivariate association with the outcome where variables with P values< 0.25 were included. The adequacy of the models in predicting the outcome variables was checked via the Hosmer–Lem test and was 0.07. The strength of the association between the different associated factors and the study outcomes was reported via crude and adjusted odd ratios, and the presence of a statistically significant association was considered at a p value less than 0.05.

## Results

### Sociodemographic characteristics of the study participants

In this study, 241 participants were involved, resulting in a response rate of 96.4%. The majority of the mothers were in the 20--29 years age group, with a mean and SD age of 28.7±4.19 years, respectively. Forty-nine percent of the children were 13–18 months of age, and 69.7% of the participants had 2–4 children. Almost eighty-five percent of the participants were from urban settings, and 43.6% of the mothers were house wives. Four-fifths percent of the mothers were married, and 23.7% of them were unable to read and write. (Table 2)

**Table 2.** The maternal and child sociodemographic characteristics of the children born to HIV positive mother on ART at Gambella Public Hospitals, South West Ethiopia, 2024.

| Variables |  | Frequency | Percent |
| --- | --- | --- | --- |
| Age in years | 20-29 | 143 | 59.3 |
|  | 30-39 | 98 | 40.7 |
| Children age in month | ≤6 | 50 | 20.7 |
|  | 7-12 | 73 | 30.3 |
|  | 13-18 | 118 | 49 |
| Sex of Children | Male | 134 | 55.6 |
|  | Female | 107 | 44.4 |
| Number of children | 1 | 42 | 17.4 |
|  | 2-4 | 168 | 69.7 |
|  | ≥5 | 31 | 12.9 |
| Residency | Urban | 204 | 84.6 |
|  | Rural | 37 | 15.4 |
| Occupation of Mother | Unemployed | 25 | 10.4 |
|  | House Wife | 105 | 43.6 |
|  | Self employed | 57 | 23.7 |
|  | Merchant | 17 | 7.1 |
|  | Daily labor | 21 | 8.7 |
|  | Other | 16 | 6.6 |
| Marital status | Never married | 11 | 4.6 |
|  | Married | 192 | 79.7 |
|  | Divorced | 36 | 14.9 |
|  | Widowed | 2 | 0.8 |
| Mother education level | Unable to read | 57 | 23.7 |
|  | Primary | 93 | 38.6 |
|  | Secondary | 64 | 26.6 |
|  | College and above | 27 | 11.2 |
| Partner education level<br>(n=192) | unable to read and write | 30 | 15.6 |
|  | primary school | 23 | 12.0 |
|  | secondary school | 50 | 26.0 |
|  | college and above | 89 | 46.4 |

### Health facility-related characteristics of the study participants

Half of the participants were 1–5 km from home to the health facility, and 90.9% of the participants did not receive necessary treatment in the health facility (Table 3).

**Table 3.** Health Facility-Related Characteristics of the Study Participants at Gambella Public Hospitals, Southwest Ethiopia, 2024.

| variable | category | frequency | Percent |
| --- | --- | --- | --- |
| Distance of health facility from home | Within 1km | 36 | 14.9 |
|  | 1-5km | 122 | 50.6 |
|  | >5km | 83 | 34.4 |
| The necessary treatment was available in all visits in your health facility | Yes | 219 | 90.9 |
|  | No | 3 | 1.2 |
|  | Not sure | 19 | 7.9 |

### Social support-related characteristics of the study participants

The majority (77.6%) of the participants had information about MTMS, and 40.7% participated in MTMS. Sixty-four percent of the partners were providing support for their children, and 95.9% were delivered at health facilities. (Table 4)

**Table 4.** Social Support-Related Characteristics of the Study Participants at Gambella Public Hospitals, Southwest Ethiopia, 2024.

| variable |  | frequency | Percent |
| --- | --- | --- | --- |
| Information about MTMS | Yes | 187 | 77.6 |
|  | no | 54 | 22.4 |
| MTMS participation | Yes | 98 | 40.7 |
|  | No | 143 | 59.3 |
| Partner support for children | Always | 45 | 18.7 |
|  | sometimes | 155 | 64.3 |
|  | never | 41 | 17.0 |
| Place of delivery | Health facility | 231 | 95.9 |
|  | home | 10 | 4.1 |

### Comorbidities and ART-related characteristics of the study participants

Almost sixteen percent of the mothers were syphilis positive, and 27.8% of the mothers were malnourished according to the MUAC measurement. Three-fourths of the study participants initiated ART before pregnancy, 19.1% initiated it during pregnancy, and 4.6% initiated it after pregnancy. Forty percent of the participants were taking ART for a duration of 6 months to 2 years, and 56.3% of the participants had good adherence to ART. (Table 5)

**Table 5.**
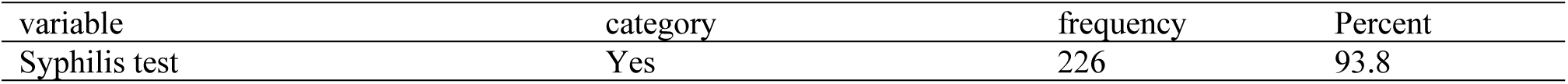
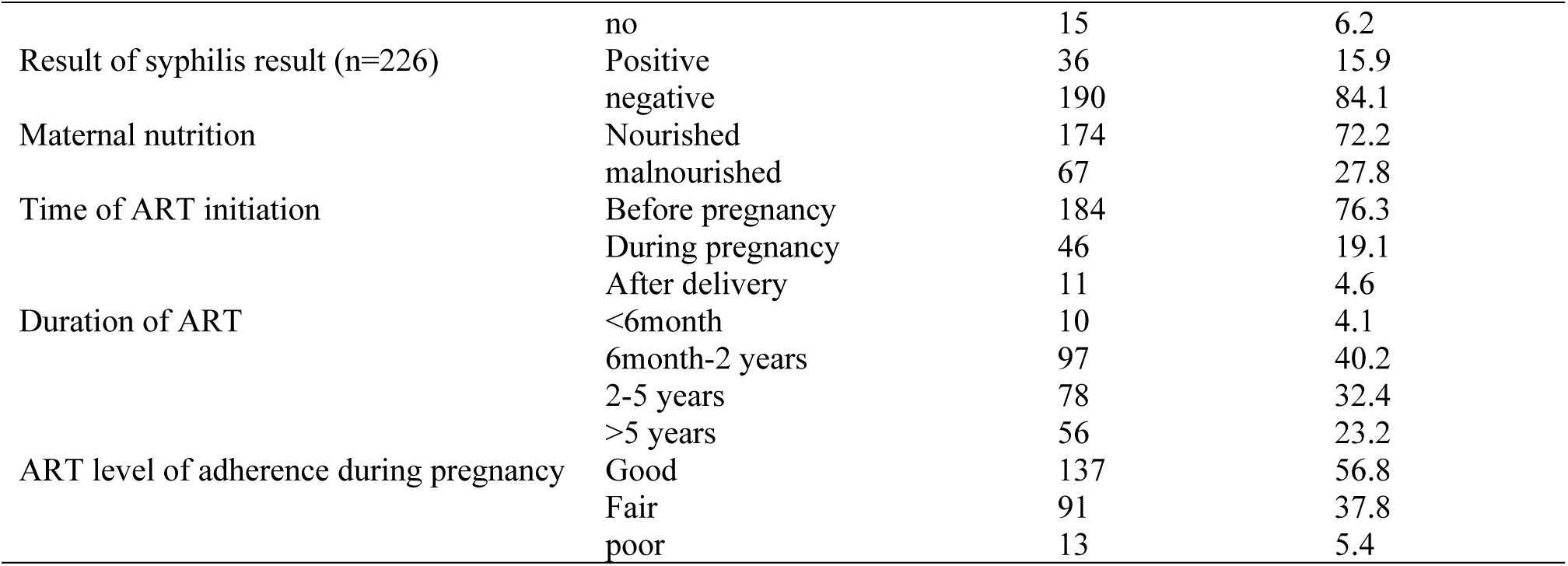
Comorbidity and ART-Related Characteristics of the Study Participants at Gambella Public Hospitals, Southwest Ethiopia, 2024.

### Obstetric characteristics of the study participants

Sixty-six percent of the study participants were multiparous, 56.8% of the women had a planned pregnancy of the current child, and 95.4% had ANC during pregnancy. Ninety-five percent of the participants delivered via a health facility, and 97.1% delivered via spontaneous vaginal delivery. (Table 6)

**Table 6:** Obstetric-Related Characteristics of the Study Participants at Gambella Public Hospitals, Southwest Ethiopia, 2024.

| variable | category | frequency | Percent |
| --- | --- | --- | --- |
| gravidity | One | 30 | 12.4 |
|  | two-four | 160 | 66.4 |
|  | Five and above | 51 | 21.2 |
| The current child pregnancy was planned | Yes | 137 | 56.8 |
|  | no | 104 | 43.2 |
| Antenatal care in the child pregnancy | Yes | 230 | 95.4 |
|  | no | 11 | 4.6 |
| Number of ANC | <4 | 87 | 37.8 |
|  | ≥4 | 143 | 62.2 |
| Place of delivery | Health facility | 229 | 95 |
|  | Home | 11 | 4.6 |
|  | Others | 1 | 0.4 |
| Mode of delivery | SVD | 234 | 97.1 |
|  | CS | 7 | 2.9 |
| Presence of episiotomy | Yes | 22 | 9.1 |
|  | no | 219 | 90.9 |

### HIV and health condition-related characteristics of the children

The findings of the study revealed that the prevalence of HIV infection among children born to HIV-positive mothers was 7% (95% CI: (4, 10)). (Figure 3)

Ninety-three percent of the children born to HIV-positive mothers received prophylaxis, and 96.6% initiated prophylaxis 24 hours after delivery. Ninety-five percent of the participants’ child feeding practices were EBF, and 59.3% of the children had good adherence to ART prophylaxis. Almost eighty-three percent of the children had normal birth weights. (Table 7)

**Table 7.** HIV and Health Condition-Related Characteristics of the Children Born to HIV Positive Mothers on ART at Gambella Public Hospitals, Southwest Ethiopia, 2024.

| variable | category | frequency | Percent |
| --- | --- | --- | --- |
| Did the child take HIV prophylaxis | Yes | 223 | 92.5 |
|  | no | 18 | 7.5 |
| ART prophylaxis initiation time after delivery (n=230) | With in 24 hrs. | 222 | 96.6 |
|  | 24-48hrs | 4 | 1.7 |
|  | After 48hrs | 4 | 1.7 |
| Feeding status | EBF | 225 | 94.9 |
|  | Mixed feeding | 9 | 3.8 |
|  | Exclusive formula feeding | 3 | 1.3 |
| Adherence to ART prophylaxis | Good | 143 | 59.3 |
|  | poor | 98 | 40.7 |
| Children birth weight | <2500 | 10 | 4.1 |
|  | 2500-3999 | 199 | 82.6 |
|  | ≥4000 | 32 | 13.3 |
| DBS result | Positive | 17 | 7.1 |
|  | negative | 224 | 92.9 |
| Time of infant starting cotrimoxazole | Before 6weeks | 9 | 3.7 |
|  | After 6 weeks | 221 | 91.7 |
|  | Not on CPT | 11 | 4.6 |

### Associated Factors of HIV Infection in Children Born with HIV-Positive Mothers on ART at Gambella Public Hospitals, Southwest Ethiopia, 2024

The associations between children’s HIV infection status and independent variables were measured by odds ratios (ORs) and 95% confidence intervals (CIs). Accordingly, nine variables were significantly associated with HIV infection in the bivariate logistic regression analysis. The multivariate logistic regression analysis revealed that children of mothers on ART aged 30--39 years were 8.7 times more likely to acquire HIV infection than were those of mothers aged 20--29 years (AOR=8.7, 95% CI=1.51--50.56). Compared with those from urban areas, children from rural areas were 6.7 times more likely to have HIV infection (AOR=6.7, 95% CI=1.38, 32.58). Children with syphilis-positive mothers were 19.6 times more likely to have HIV infection than those with syphilis-negative mothers were (AOR=19.6, 95% CI=3.09, 44.43). Additionally, children from unplanned pregnancies were 10.4 times more likely to have HIV infection than those from planned pregnancies were (AOR=10.4, 95% CI=1.45, 75.29). (Table 8)

**Table 8.** Bivariate and Multi variable Logistic Regression of the Association Between Independent Variables and HIV Infection Among Children Born to HIV Positive Mothers in Gambella Public Hospitals, Southwest Ethiopia, 2024.

| Variable | HIV exposed children<br>HIV status |  | COR with 95%CI | P-value | AOR with 95%CI |
| --- | --- | --- | --- | --- | --- |
|  | positive | negative |  |  |  |
| Maternal age |  |  |  |  |  |
| 20-29 | 2 | 141 |  | 1 |  |
| 30-39 | 15 | 83 | 12.7(2.84, 57.11) | 0.015 | **8.7(1.51, 50.56)** |
| Residency |  |  |  |  |  |
| Urban | 8 | 196 |  | 1 |  |
| Rural | 9 | 28 | 7.9(2.81, 22.09) | 0.018 | **6.7(1.38, 32.58)** |
| Information about MTMS |  |  |  |  |  |
| Yes | 10 | 177 |  | 1 |  |
| No | 7 | 47 | 2.6(0.95, 7.29) | 0.211 | 3.2(0.52, 19.47) |
| Syphilis result |  |  |  |  |  |
| Positive | 10 | 26 | 10.1(3.52, 28.72) | 0.002 | **19.6(3.09, 44.43)** |
| Negative | 7 | 183 |  | 1 |  |
| Maternal nutrition status |  |  |  |  |  |
| Nourished | 10 | 164 |  | 1 |  |
| Malnourished | 7 | 60 | 1.9(0.69, 5.25) | 0.349 | 2.3(0.40, 13.39) |
| Know HIV status prior to pregnancy |  |  |  |  |  |
| Yes | 11 | 210 |  | 1 |  |
| No | 6 | 14 | 8.2(2.64, 25.39) | 0.863 | 1.4(0.02, 92.33) |
| Planned pregnancy |  |  |  |  |  |
| Yes | 2 | 135 |  | 1 |  |
| No | 15 | 89 | 11.4(2.54, 50.96) | 0.020 | **10.4(1.45, 75.29)** |
| ANC |  |  |  |  |  |
| Yes | 13 | 217 |  | 1 |  |
| No | 4 | 7 | 9.5(2.47, 36.79) | 0.399 | 7.4(0.07, 78.66) |
| Child prophylaxis adherence |  |  |  |  |  |
| Good | 7 | 136 |  | 1 |  |
| Poor | 10 | 88 | 2.2(0.81, 6.02) | 0.313 | 10.4(1.45, 75.29) |

## Discussion

The findings of this study revealed that the prevalence of HIV infection among children born to HIV-positive mothers was 7% (95% CI: 4, 10). This finding was lower than that of a study performed at Kisii Teaching and Referral Hospital, Kenya (13.5%) (24), and a study performed in the Amhara region (10.1%) (22). This difference might be due to differences in study design, sample size and study area. In addition, the difference might be due to health care-seeking behavior.

This finding is also in line with studies performed in districts such as Karnataka, India (7.8%) (25); systematic reviews and meta-analyses (7.68%) (26); studies performed in Bahrdar, Ethiopia (6.8%) (27); the University of Gondar Specialized Hospital (5.5%) (28); Northwest Ethiopia (6.1%) (29); and studies conducted in the Sidama region (9%) (30). This similarity might be due to the implementation of a similar program to the PMTCT program.

On the other hand, this finding was significantly greater than that reported in a study performed in Ethiopia (2.6%) (31). This difference may be due to differences in sample size.

Compared with mothers aged 20--29 years, mothers aged 30--39 years had a greater risk of HIV infection. This finding was supported by studies performed in Belgaum district, Karnataka, India (25), in Ethiopia (31). This may be because as women age, their immune system naturally weakens, which may affect how well they control HIV. This increased viral load heightens the risk of mother-to-child transmission during pregnancy, childbirth, or breastfeeding. Older mothers are more likely to have comorbid conditions such as hypertension, diabetes, or other chronic diseases that may complicate pregnancy and the management of HIV. Some of these conditions can lead to increased viral activity or complicate ART use, contributing to a higher risk of MTCT.

Children whose parents were living in rural areas had a greater risk of HIV infection than did those whose parents were living in urban areas. This finding was congruent with that of a study performed in southern Ethiopia (32). This may be because rural areas often have fewer healthcare facilities, and mothers may have difficulty accessing regular antenatal care (ANC). This can lead to missed opportunities for early HIV testing, timely initiation of antiretroviral therapy (ART), and proper monitoring during pregnancy.

Children whose mothers were syphilis positive were more likely to have HIV infection than those whose mothers were syphilis negative were. This finding was in line with a study performed in Addis Ababa, Ethiopia (33). This may be because syphilis infection in the mother can cause an inflammatory response in the body, which can increase the viral load of HIV. Higher HIV viral loads are strongly associated with an increased risk of mother-to-child transmission during pregnancy, childbirth, and breastfeeding.

Pregnant children who were unplanned had higher odds of acquiring HIV infection than did those in the opposite compartment. This finding was supported by a study performed in Addis Ababa, Ethiopia (33). This may be because unplanned pregnancies may lead to delays in starting ART if the mother discovers her pregnancy late or does not access healthcare services promptly. Women with unplanned pregnancies may face challenges in adhering to ART regimens due to emotional stress or other unpreparedness, which can lead to higher viral loads and an increased risk of HIV transmission to the baby.

## Conclusion

The study revealed that 7% of children born to HIV-positive mothers were infected with HIV. While Ethiopia has made significant progress in reducing mother‒child transmission (MTCT) over the last decade, the rate reported in this study is still relatively high.

The factors associated with HIV infection in children were mothers aged 30--39 years, children living in rural areas, children who were positive for syphilis, and pregnant children with unplanned HIV infection.

## Data Availability

The dataset supporting the findings of this study is available upon reasonable request. Due to ethical considerations and the need to protect participant confidentiality, access is restricted and may be granted to researchers who meet the criteria set by the Ethical Review Committee of the College of Public Health at Mettu University.

## Declarations

### Compliance with ethical standards

This study was conducted in accordance with the ethical principles outlined in the Declaration of Helsinki. Ethical approval was obtained from the institutional review board (IRB) of the Ethical Review Committee of the College of Public Health at Mettu University. Written informed consent was obtained from each of the study participants after their caregivers were informed about the nature and objectives of the study. The participants were informed of their right to withdraw from the study at any time without any repercussions on the quality of care they received. To ensure confidentiality, personal identifiers such as names were not included in the questionnaires. The data collected were used solely for the purposes of this study and were kept secure in a locked location accessible only to the principal investigator.

### Consent for Publication

All authors have given their consent for the publication of this manuscript. They affirm that the final version of the manuscript has been read and approved by all authors, and they consent to its publication. They agree to be accountable for all aspects of the work, ensuring that questions related to the accuracy or integrity of any part of the work are appropriately investigated and resolved.

### Availability of Data and Materials

Data will be available upon the request of the corresponding author.

## Acknowledgment

I would like to thank Gambella General Hospital, Gambella primary Hospital, Pugudo Primary Hospital and the department of public health at Mettu University, college of health sciences, for your support in this research.

